# Diabetes mellitus is associated with a shared hyper-inflammatory immune response in melioidosis and tuberculosis patients: an observational case-control study

**DOI:** 10.1101/2024.05.31.24308267

**Authors:** Patpong Rongkard, Barbara Kronsteiner, Clare Eckold, Kemajittra Jenjaroen, Suchintana Chumseng, Parinya Chamnan, Mohammad Ali, Emanuele Marchi, Direk Limmathurotsakul, Narisara Chantratita, T. Eoin West, Sina A. Gharib, Jacqueline M. Cliff, Nicholas P.J. Day, Paul Klenerman, Susanna J. Dunachie

**Affiliations:** NDM Centre for Global Health Research, Nuffield Department of Clinical Medicine, University of Oxford, Oxford, UK; Mahidol-Oxford Tropical Medicine Research Unit, Mahidol University, Bangkok, Thailand; Tuberculosis Centre and Department of Infection and Biology, London School of Hygiene and Tropical Medicine, London, UK; Sunprasithiprasong Hospital, Ubon Ratchathani, Thailand; Peter Medawar Building for Pathogen Research, Nuffield Department of Clinical Medicine, University of Oxford, Oxford, UK; Department of Microbiology, Mahidol University, Bangkok, Thailand; Division of Pulmonary, Critical Care and Sleep Medicine, University of Washington, USA; Department of Global Health, University of Washington, USA; Translational Gastroenterology and Liver Unit, Nuffield Department of Clinical Medicine, University of Oxford, UK; NIHR Oxford Biomedical Research Centre, Oxford University Hospitals NHS Foundation Trust, Oxford, UK; Department of Biochemistry and Molecular Biology, Bangabandhu Sheikh Mujib Medical University, Shahbag, Dhaka, Bangladesh

## Abstract

**Background:** Melioidosis is a serious infection caused by the bacterium *Burkholderia pseudomallei* with a case fatality rate of up to 40% in Northeast Thailand. Diabetes increases the risk of developing melioidosis by 12-fold. A similar, but less marked, relationship with diabetes is seen in tuberculosis (TB) patients, with a 3-fold increased risk of developing TB in people with diabetes. However, the mechanisms underlying increased susceptibility are not fully understood.

**Methods:** 81 acute melioidosis patients from Northeast Thailand and 151 TB patients from South Africa, Indonesia, Romania and Peru alongside uninfected control cohorts were studied by whole blood RNA sequencing. Both supervised and unsupervised data analysis approaches were performed including differential gene expression (DGE) analysis, pathway analyses, and weighted gene co-expression network analysis (WGCNA).

**Results:** Diabetes status was associated with a hyper-inflammatory response to both melioidosis and TB, with increased neutrophil and platelet degranulation, and exaggerated activation of coagulation and scavenger activation pathways, alongside decreased phosphoinositide-3-kinase protein kinase B (P13K-Akt) signalling. In melioidosis, changes with diabetes were subtle but also included increased tumour necrosis factor (TNF) signalling via nuclear factor kappa-light-chain-enhancer of activated B cells (NFκB) and enhancement of endoplasmic reticulum stress and unfolded protein responses. Diabetes-related changes were more distinct in TB, with marked reduction of interferon signalling responses.

**Conclusion:** Diabetes is associated with enhanced non-specific inflammatory responses in both melioidosis and TB and an impaired interferon-mediated response to TB, with implications for future host-directed therapies.

**Summary:** Diabetes status is associated with a hyper-inflammatory response to both melioidosis and TB, with increased neutrophil and platelet degranulation, and exaggerated activation of coagulation and scavenger activation pathways. The impact is more subtle in melioidosis but pronounced in tuberculosis with stunted interferon responses.

## Introduction

Melioidosis is a deadly infectious disease caused by infection with *Burkholderia pseudomallei* (Bp), a Gram-negative bacillus with estimated 165,000 cases and 89,000 deaths globally each year (*1*). The case fatality rate for people admitted to hospital with melioidosis is up to 40% in Northeast Thailand and 14% in Australia (*2, 3*). Risk factors include chronic renal disease, excessive alcohol consumption, older age and diabetes mellitus (DM), which carries a 12-fold increased risk (*2, 4*).

*Mycobacterium tuberculosis* (Mtb) causes tuberculosis (TB) and is the leading cause of death from a single pathogen worldwide (TB) (*5*). In 2021, around 10 million people were infected with Mtb, with 1.3 million deaths (*5*). Risk factors for developing TB disease are HIV infection, undernourishment, alcohol abuse, smoking, and DM (*6, 7*). DM is associated with up to a three-fold increased risk of developing TB (*7, 8*) and contributed to approximately one million TB cases worldwide in 2013 (*9*). Melioidosis and TB share some common features, with both diseases caused by intracellular bacteria, have lung disease as the commonest clinical presentation, and feature granuloma formation (*10*). Type I immune responses are important for controlling both diseases (*11, 12*). Interestingly, HIV infection is not a major risk factor for acquiring melioidosis (*13*).

Multiple host immune response pathways are compromised in people with DM including phagocytosis, cytokines, and chemokine response (*14*). However common pathways underlying the specific susceptibility to melioidosis and TB in patients with DM have not been well characterised. We hypothesised that the increased risk in patients with DM to melioidosis and TB is underlined by common transcriptomic profiles during infection. In this study, we examined the relationship between whole blood transcriptomic profiles associated with DM during melioidosis and TB.

## Materials and methods

### I. Study design and ethical approval

For the melioidosis cohort, we conducted a prospective observational study (MICRO1501) between 2015 and 2017 at Sunpasitthiprasong Hospital, Ubon Ratchathani, Thailand. The study protocol was approved by the ethics committees of the Faculty of Tropical Medicine, Mahidol University (TMEC 12-014); Sunpasitthiprasong Hospital, Ubon Ratchathani (017/2559) and the Oxford Tropical Research Ethics Committee (OXTREC35-15). The study was conducted according to Good Clinical Practice, and all subjects gave written informed consent, including for export and storage of their blood samples. We recruited adults aged 18 years and over into the Melioidosis Cohort of the study as soon as feasible after hospitalisation with melioidosis, defined as culture of Bp from any clinical specimen. We also recruited healthy household contacts of melioidosis cases enrolled in the study as endemic control participants to minimise demographic bias between cases and controls, alongside people with DM recruited from the DM out-patient clinic at Sunpasitthiprasong hospital. We measured HbA1c for all participants and defined diabetes status for the Melioidosis Cohort as holding a pre-existing diagnosis of DM and / or having an HbA1c ≥ to 6.5 %, according to WHO criteria (*15, 16*).

For the TB cohort, we used whole blood bulk RNA sequencing data from a previously published study (*16, 17*). This study included 239 adults participants aged 18 years or older with newly diagnosed, bacteriologically confirmed pulmonary TB with and without DM along with uninfected patients with DM and healthy donors between 2013 and 2016 across four-study sites: South Africa, Indonesia, Romania and Peru (*17*). The TB patients were further classified as 1) “TB-only”: TB without DM (HbA1c level < 5.7%), 2) “TB-IH”: TB with intermediate hyperglycaemia or IH (HbA1c level between ≥5.7 and < 6.5%), and 3) “TB-DM”: TB with DM (HbA1c level between ≥ 6.5%). The study size was selected based on samples available and in line with successful studies yielding findings of interest in the peer-reviewed literature.

#### Sample collection

For the melioidosis cohort, we collected 3 mL whole blood samples into Tempus blood RNA tube (Applied Biosystems) from culture-confirmed melioidosis (Melioidosis Cohort, n=81) along with control cohorts, DM outpatients (DM, n=15), and household contacts of the melioidosis patients (HH, n=14). The tempus blood RNA tubes were store at -80°C until further processing.

### II. RNA sequencing and data acquisition

For the melioidosis cohort, we performed RNA isolation and preparation according to the manufacturer’s instructions unless otherwise stated. We isolated total RNA from whole blood sample collected in Tempus blood RNA tube using the Tempus Spin RNA Isolation (Applied Biosystems, Thermo Fisher Scientific). We performed library preparation and RNA sequencing at the Oxford Genomics Centre, Wellcome Centre for Human Genetics, with ribozero library preparation and the globin depletion workflow and we sequenced the library for 75-base pair pair-end, 50 million reads using a HiSeq4000 sequencer. For the TB cohort, *Eckold et al* (*16*) generated the expression data and provided the raw sequencing data (FASTQ files). Briefly, whole blood was collected into PAXgene Blood RNA Tubes (PreAnalytiX, Qiagen) and sequenced from the PolyA tail library preparation approach. For this study we used the same upstream data analysis pipeline including quality check, mapping, and read counting for both melioidosis and TB cohorts (Supplemental Information).

### III. Statistical Analysis

Expression data (transcripts) were annotated to gene symbol. Genes with no counts or lowly expressed were removed. An overview of data analysis pipeline is shown in Figure 1A. Briefly, differential gene expression (DGE) analysis was carried out using negative binomial generalised linear model implemented in DESeq2 R package (version 1.34.0) adjusted for age and sex covariates (*18*). Following DGE analysis, functional pathway analysis was performed using clusterProfiler R package (version 4.6.2) (*19*). Gene set enrichment analysis (GSEA) based on the MSigDB gene sets was performed using fgsea R package (version 1.18.0) (*20*). To identify co-expressed gene modules associated with DM and corresponding hub genes during melioidosis and TB, we performed weighted gene co-expression network analysis (WGCNA) (*21*). Subsequently, co-expressed modules that displayed significant relationship to clinical variables were subjected to pathway enrichment analysis. Lastly, to identify immune cell populations associated with DM, cell type abundances were estimated from bulk RNA sequencing data using xCell (version 1.1.0) (*22*).

**Figure 1.**
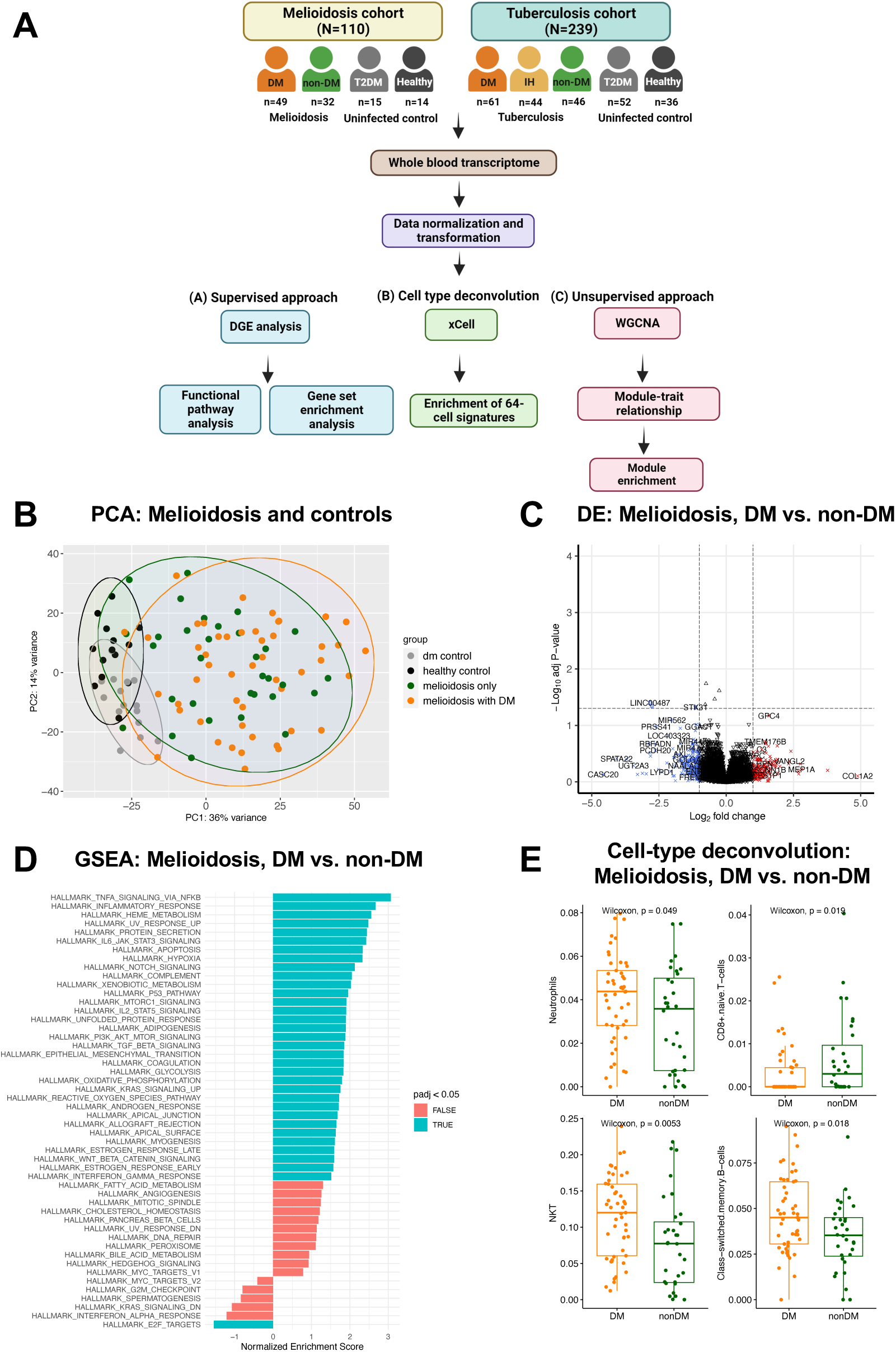
Data analysis pipeline and whole blood transcriptomic profiling in acute melioidosis patients with diabetes. (A) Core data analysis approaches for both melioidosis and tuberculosis study cohorts consist of supervised approach, cell type deconvolution, and unsupervised approach. (B) Principal component analysis (PCA) of the top 1,000 most variable genes among melioidosis patients and control cohorts. Melioidosis cohort is divided into the patients with diabetes (“melioidosis with dm”, orange dots, n=49), without diabetes (“melioidosis only”, green dots, n=32), uninfected diabetes outpatients (“dm control”, grey dots, n=15), and uninfected healthy donors (“healthy control”, grey dots, n=14). (C) Volcano plot of differentially expressed genes between melioidosis patients with diabetes and without diabetes. Differentially expressed genes were based on absolute (Log_2_ fold-change)≥ 1 (x-axis) and adjusted P-value < 0.05 (y-axis) (dotted lines). (D) Gene set enrichment analysis between melioidosis patients with diabetes and without diabetes based on Hallmark gene sets. Normalised enrichment scores are displayed where significant pathways were labeled in turquoise (false discovery rate (FDR) <0.05). (E) Significant immune cell-subset enrichment between melioidosis patients with diabetes and without diabetes. The statistical analysis was performed using Mann-Whitney test, and the corresponding P-value was displayed on each plot along with median and inter-quartile range boxes.

## Results

### Melioidosis causes profound changes in the whole blood transcriptome

The Melioidosis cohort consists of 81 patients with culture-proven melioidosis, in which majority of the patients (49, 61%) had DM by the time of enrolment (Table 1), alongside 29 healthy controls with and without DM (Figure 1A). We sampled the patients a median of 5 days (IQR 4-6 days) after admission to hospital. The case fatality rate was 35% with no difference between DM and non-DM. To determine whether melioidosis causes changes in gene expression, we performed principal component analysis (PCA) on the 1,000 most variable genes in melioidosis and control cohorts. PCA shows clear separation between melioidosis patients and control groups (Figure 1B). There was also some separation between healthy controls and patients with DM. However, no clear separation was observed by DM status within the melioidosis patients. Next, we compared gene expression between healthy controls and patients with DM by DGE and identified 94-upregulated and 2-downregulated genes in DM patients compared to healthy controls (absolute [log2 fold change] ≥1, adjusted P-value <0.05) (Supplementary Figure 1A). Thus, healthy controls were used as the baseline for subsequent analyses. DGE analysis identified large-scale changes in gene expression between melioidosis patients and healthy donors (Supplementary Figure 1B). Pathways involved in inflammatory immune responses such as inflammatory response, TNF signalling via NF-κB, neutrophil and platelet degranulation were highly up-regulated in melioidosis compared to healthy control (Supplementary Figure 1C).

**Table 1.** Demographics of melioidosis study cohort. Subject demographics for whole blood transcriptomic study from melioidosis and control cohort

| Baseline characteristics | Melioidosis with diabetes (n=49) | Melioidosis only (n=32) | Healthy control (n = 14) | Diabetes control (n = 15) | P values |
| --- | --- | --- | --- | --- | --- |
| Female, no. (%) | 20 (41%) | 7 (22%) | 6 (43) | 8 (53) | †0.07 |
| Age in years; median (IQR) | 55 (49-63) | 65 (52-73) | 47 (44-54) | 49 (44-57) | ‡0.01 |
| Non-survivors*, no. (%) | 23 (47%) | 13 (41%) | NA | NA | †0.58 |

Diabetes (DM) is defined by those patients who were previously diagnosed with diabetes or having an HbA<sub>1c</sub> level $\geq 6.5\%$ on recruitment into the study. \*Case fatalities are based on 28-day mortality. † Chi-square test between melioidosis with and without diabetes. ‡ Mann-Whitney test between melioidosis with and without diabetes. NA= Not applicable.

**Table 2.**
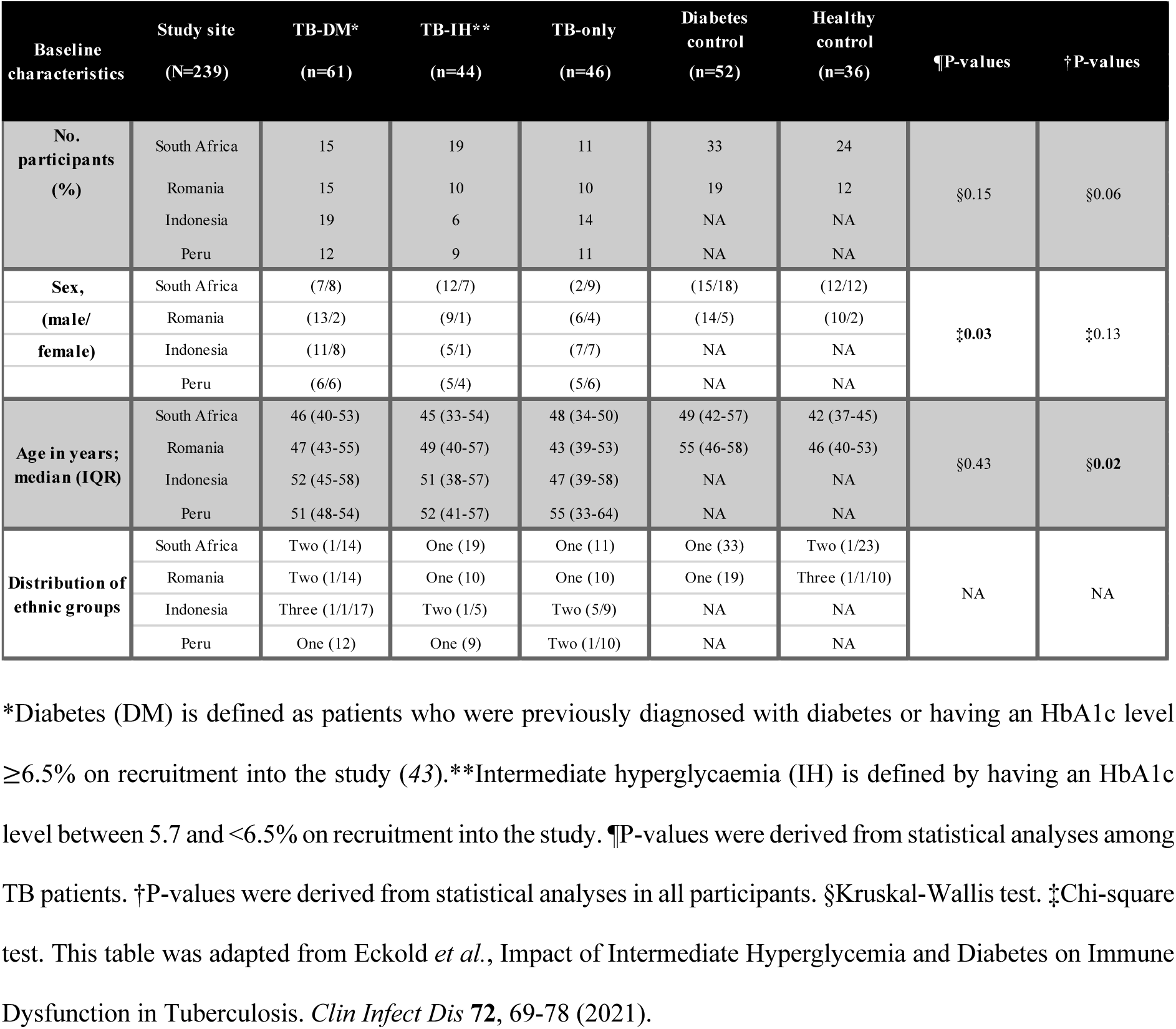
Subject demographics for tuberculosis study cohorts. Subject demographics for whole blood transcriptomic study from tuberculosis and control cohort

### Melioidosis patients with diabetes have an increased inflammatory immune response and cellular stress response

DGE analysis did not reveal substantial differences between melioidosis patients with and without DM (Figure 1C). We next performed genome-wide functional class scoring using pre-ranked gene set enrichment analysis (GSEA) following DGE analysis. GSEA by Hallmark gene sets revealed enrichment in melioidosis patients with DM of pathways involved in inflammation and cellular stress responses, such as TNF signalling via NF-κB, protein secretion, and heme metabolism (Figure 1D). Cell-type deconvolution analysis demonstrated that melioidosis patients with DM have an increased number of natural killer T (NKT) cells, class-switched memory B cells, and neutrophils, but fewer naive CD8+ T cells compared to those without DM (Figure 1E).

### Co-expressed gene modules enriched for inflammatory immune responses and cellular stress response are associated with diabetes during melioidosis

To complement the results obtained from DGE and pathway analyses in an unsupervised manner, we performed WGCNA. 22 co-expressed gene modules (module eigengenes, MEs – each identified by a colour) were identified (Figure 2B). Module-trait relationship analysis identified significant correlations between DM status clinical trait and ME salmon (Pearson’s rho=0.25, P-value=0.03) and ME blue (Pearson’s rho = 0.23, P-value=0.04) (Figure 2C). Pathway analysis identified enrichment of pathways involved in pro-inflammatory immune responses such as the innate immune system, neutrophil degranulation, and NOD-like receptor signalling pathway within the ME blue module. Furthermore, ME salmon – which was associated with DM status and HbA1c level identified enriched pathways involved in regulation of endoplasmic reticulum and response to unfolded protein (Figure 2D).

**Figure 2.**
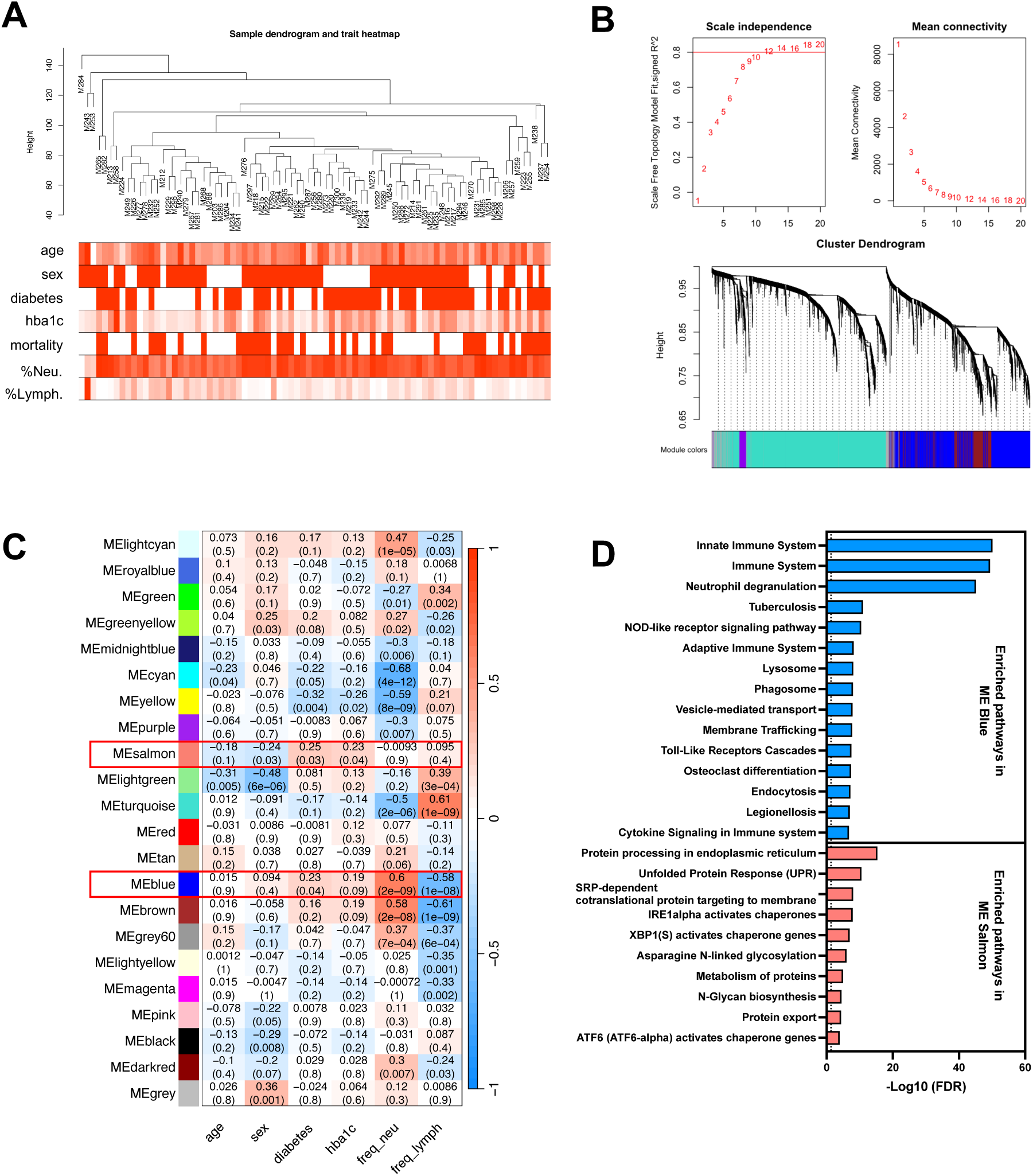
Weighted gene co-expression network analysis in melioidosis patients. Weighted gene co-expression network analysis (WGCNA) was performed in 81 melioidosis patients. (A) Sample dendrogram with corresponding clinical data. (B) Scale-free topological analysis and cluster dendrogram of co-expressed module (module eigengene, ME). (C) Module-trait relationship between ME and clinical traits. (D) Pathway enrichment analysis within MEs with significant correlation (ME Blue and ME Salmon) to diabetes status based on Reactome and KEGG gene sets. The enriched pathways are displayed by -Log_10_ (False Discovery Rate, FDR) values and dotted line defines FDR < 0.05. Clinical traits include age, sex, diabetes status (diabetes), HbA_1c_ level (hba1c), %neutrophils (freq_neu), and %lymphocytes (freq_lymph).

### Hyperglycaemia and diabetes drive an increased inflammatory immune response in both melioidosis and TB patients

To compare the impact of DM and intermediate hyperglycaemia on the whole blood transcriptomic of both TB and melioidosis (Figure 3), we first undertook novel analysis of a previously published transcriptomics study in TB (*16*). Functional pathway analysis was employed following DGE analysis (absolute [Log2 fold-change]≥1, adjusted P-value < 0.05) by Reactome and KEGG gene sets (Supplementary Figure 2 and 3). Pathways involved in type I immune responses such as interferon signalling, interferon alpha/beta signalling and interferon gamma signalling were upregulated in all TB groups compared to healthy controls (Figure 3A and Supplementary Figure 4A). However, anti-viral mechanisms by IFN-stimulated genes, ISG15 antiviral mechanisms, hepatitis C, measles, and creation of C4 and C2 activators pathways were exclusively up-regulated in the TB-only group. Similarly, GSEA based on Hallmark gene sets, revealed that TB-IH patients had highly enriched pathways involved in inflammatory immune responses and metabolism such as coagulation, IL-6 JAK STAT3 signalling, complement, and reactive oxygen species, whilst, interferon alpha and gamma pathways were enriched in TB-only compared to TB-DM patients (Supplementary Figure 4C & 4D).

**Figure 3.**
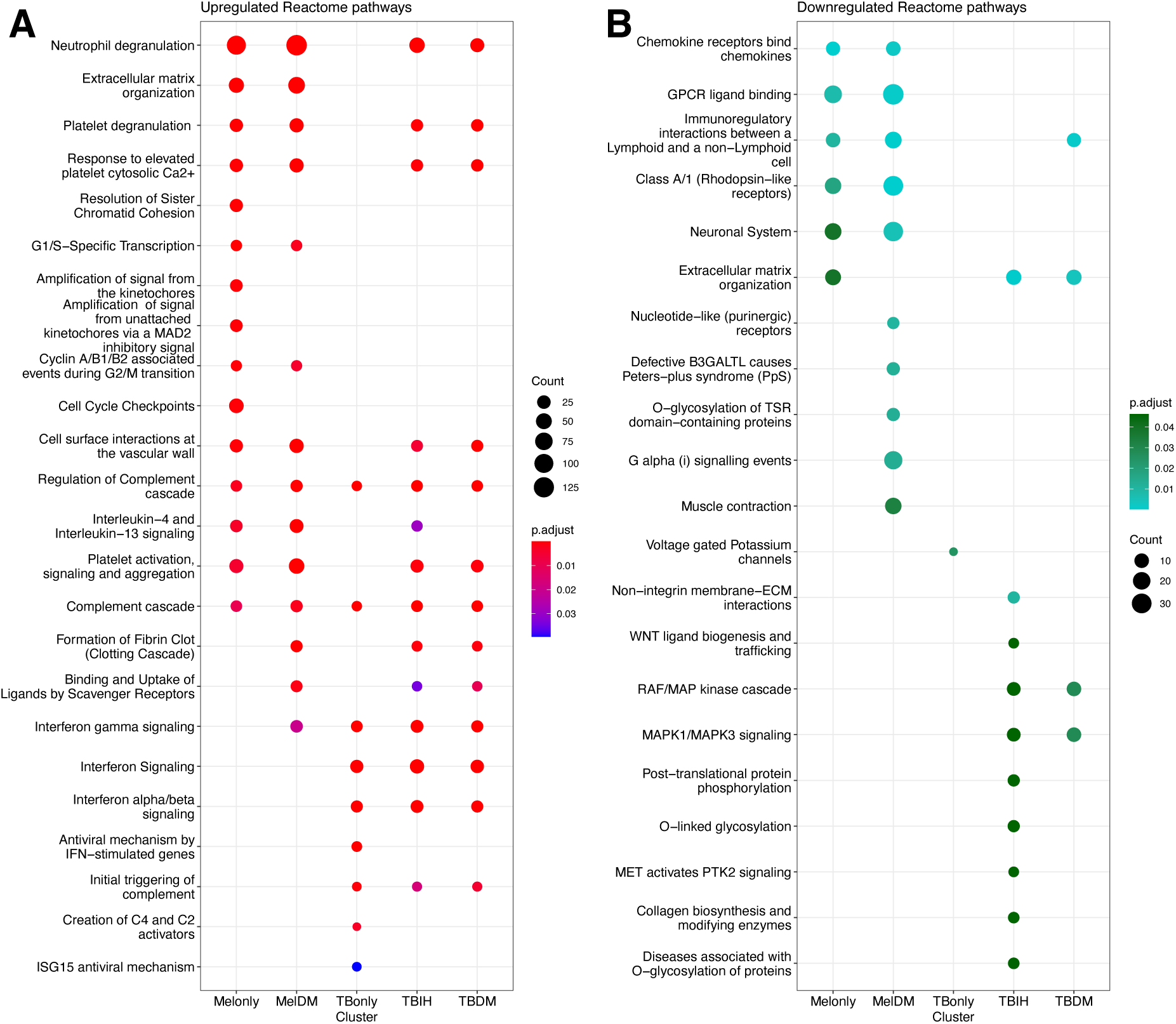
Functional pathway analysis among melioidosis and tuberculosis patients living with and without diabetes. Functional pathway analysis based on Reactome gene sets following differential gene expression (DGE) analyses among tuberculosis (TB) from South African cohort and melioidosis cohort. (A) Upregulated and (B) downregulated pathways in melioidosis and TB patients compared to their corresponding uninfected healthy control cohorts respectively. The gradient colour bar corresponds to the adjusted P-value. The size of each term is indicated by representative counts (number of DEGs). Differentially expressed genes were pre-filtered based on a cut-off of absolute[Log2 fold-change] ≥1 and adjusted P-value <0.05. Melonly and MelDM = Pathways derived from DGE analysis between melioidosis without diabetes and with diabetes compared to healthy control respectively. TBonly, TBIH, and TBDM = Pathways derived from DGE analysis among TB without diabetes, TB with intermediate hyperglycaemia, and TB with diabetes compared to healthy control respectively.

We identified common pathways with up-regulation in DM compared to no DM for both TB (TB-DM, TB-IH), and melioidosis, such as clotting cascade and scavenger receptors pathway responsible for ligand binding and uptake. For inflammation and immune response pathways related to infection, only melioidosis with DM displayed upregulation in TNF signalling, deficient type 1 immunity, and impaired intracellular killing mechanisms. Lastly, the PI3K-Akt signalling pathway was downregulated in TB-IH, TB-DM, and melioidosis with DM (Supplementary Figure 4C & 4D).

### Enriched neutrophils and inflammatory immune responses in co-expressed gene modules are associated with diabetes during tuberculosis

WGCNA was performed comparing TB-DM and TB-only cohorts to identify gene modules and hub genes that are associated with DM (Supplementary Figure 5A-D). 23 co-expressed gene modules were identified (Supplementary Figure 5D). Significant correlations were found between ME darkturquoise and HbA1c level and DM status through module-trait relationship analysis (Figure 4A). Module enrichment analysis based on Reactome and KEGG gene sets within ME darkturquoise (correlated with DM status and HbA1c level) identified enriched pathways involved in defence against infections driven by the innate immune compartment such as neutrophil degranulation, antimicrobial peptides, and alpha-defensins (Figure 4B). The top 20 hub genes were identified within ME darkturquoise (Figure 4C), of which 2 genes including *CEACAM8* and *BPI* were upregulated in TB-DM patients compared to TB-only patients (Figure 4D). Finally, there was significant enrichment of neutrophils and Th2 cells in TB-DM patients compared to TB patients without DM (Supplementary Figure 6).

**Figure 4.**
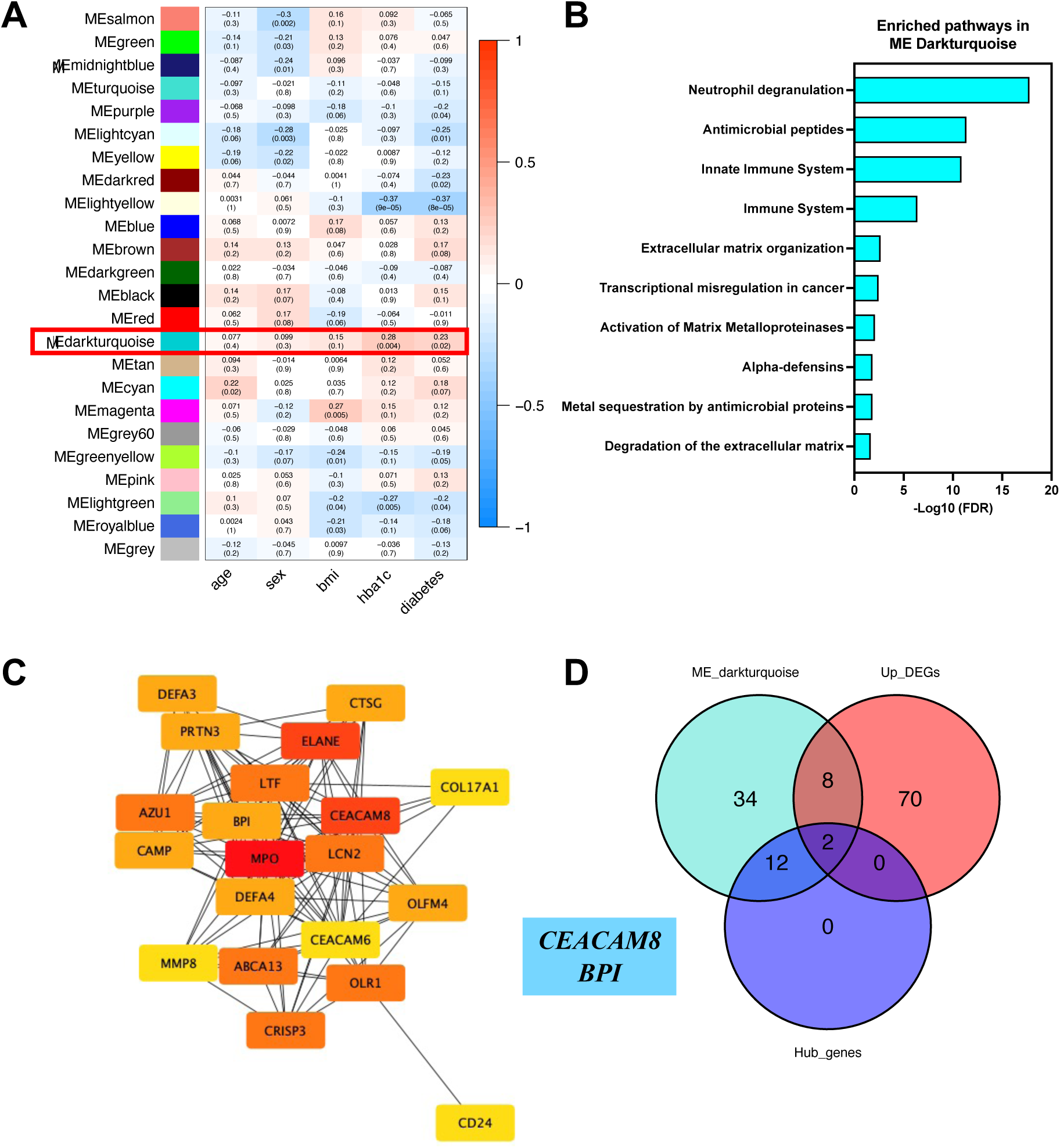
Weighted gene co-expression network analysis and identification of hub genes associated with diabetes in tuberculosis patients. (A) Module-trait relationship between co-expressed gene modules (module eigengene, ME) derived from weighted gene co-expression network analysis (WGCNA) in TB patients with and without diabetes. Clinical traits include age, sex, body mass index (bmi), HbA_1c_ level (hba1c), and diabetes status (diabetes). (B) Pathway enrichment analysis within ME with significant correlation (ME darkturquoise) to diabetes status and HbA_1c_ level based on Reactome and KEGG gene sets. The enriched pathways are displayed by -Log_10_ (False Discovery Rate, FDR) values. (C) The top 20 hub genes identified in the module eigengene (ME) darkturquoise associated with diabetes status. The hub genes were identified based on The Maximal Clique Centrality (MCC) algorithm using CytoHubba plugin on Cytoscape software. The MCC scores of hub genes were ranked from high (red) to low (yellow), and 6 hub genes without an MCC score were excluded. (D) Venn diagram was constructed to show the overlap between differentially expressed genes (DEGs), gene membership of the ME darkturquoise identified by weighted gene co-expression network analysis, and hub genes identified within the ME darkturquoise. The DEGs were identified based on up-regulated genes between TB patients with and without diabetes, using a cutoff of absolute[Log2 fold-change]≥0.5 and adjusted P-value <0.05.

## Discussion

Despite an estimated twelve-fold increased risk of melioidosis in people with DM, differences in the transcriptomic profiles between melioidosis patients with and without DM were subtle, but we were able to characterise the influences of DM using GSEA and WGCNA. Melioidosis patients with DM showed enrichment of multiple pathways involving the inflammatory immune responses and cellular stress responses by endoplasmic reticulum. These findings of enrichment of neutrophils and inflammatory responses were also identified in DM during TB.

Previous studies have demonstrated that neutrophils play a detrimental role in DM by increasing non-specific inflammatory immune responses (*23, 24*). Hyperglycaemia, a hallmark of T2DM, results in neutrophil dysfunction with impairments observed in chemotaxis, phagocytosis, and killing mechanisms (*25*). Furthermore, increased inflammation in hyperglycaemia produces microparticles under oxidative stress which stimulate the inflammasome, leading to an increased inflammatory immune response (*26*). Neutrophils under hyperglycaemic conditions are highly susceptible to NETosis contributing to inflammation and tissue damage (*27*). Overall, our study shows increased neutrophil-mediated immune response which may be contributing to disorganised immune response and detrimental inflammation.

Interferon-gamma (IFN-γ) is essential for controlling intracellular infections like TB and melioidosis (*11, 28*). In TB, CD4+ T cells are the main source of IFN-γ during the adaptive immune response against Mtb infection (*29*) hence individuals with uncontrolled HIV are particularly susceptible to the disease (*30*). However, in melioidosis, deficiencies in the CD4+ T cell response may be compensated by other immune cells, such as CD8+ T cells (*11*). This is supported by a study that found no evidence of increased susceptibility to melioidosis in HIV+ individuals (*13*). Our previous work has demonstrated reduced Bp-specific CD8+ T cell responses during melioidosis in people with DM compared to non-DM, with gamma-delta T cell responses associated with survival in DM (*31*). Overall, these findings suggest that type I immune responses are important for controlling both TB and melioidosis, but the mechanisms by which these responses are mediated may differ between the two diseases.

Our analysis confirms that the transcriptome of whole blood from TB patients is dominated by interferon signalling pathways and the complement cascade. However, TB-DM and TB-IH patients show reduced expression of interferon signalling genes and pathways, compared to TB-only patients. Additionally, TB-DM and TB-IH patients show up-regulated pathways involved in inflammation driven by the innate immune compartment. Melioidosis patients have similar transcriptomic profiles, with up-regulation of pathways involved in inflammatory immune responses. In contrast, phosphatidylinositol 3-kinase (PI3K)-Akt signalling pathway was mutually down-regulated in TB-DM, TB-IH, and melioidosis patients with DM. The PI3K-Akt signalling pathway regulates many cellular functions such as glucose metabolism, apoptosis, and immune response (*32*). Insulin resistance, hyperglycaemia, and excess free fatty acids (FFAs) have been linked to dysregulation of the PI3K-Akt signalling pathway. FFAs can inhibit activation of the insulin pathway but activate protein kinase C which triggers the NF-kB pathway (*33*). The latter pathway may play a role in increased inflammatory immune response in DM (*34*). Our findings suggest that patients with DM who are also infected with Mtb or Bp may experience excessive inflammation leading to impaired cellular immune function, potentially mediated by neutrophils and innate immune components as described earlier.

In this study, we reanalysed data from *Eckold et al.* (*16*) and further identified genes potentially playing deleterious roles in TB patients with DM. The two hub genes including *CEACAM8* and *BPI* were identified and differentially expressed in TB-DM compared to TB-only. *CEACAM8* encodes a cell adhesion module (CD66b) which is highly expressed on neutrophils during activation (*35*) and is linked to increased disease severity and poor outcome in inflammatory diseases and acute infections (*36, 37*). Several studies have demonstrated that expression of CD11b on monocytes, together with CD66b on neutrophils, are increased in DM patients compared to healthy donors (*38, 39*). The increased cell adhesion molecules on neutrophils may be associated with increased risk of developing DM-related complications such as atherosclerosis (*38*). *BPI* encodes Bactericidal Permeability Increasing Protein (BPI) which is found in the azurophilic granules of neutrophils and has binding specificity and neutralisation activity against lipopolysaccharide (LPS) from Gram-negative bacteria (*40*). In TB, BPI can recognise lipoarabinomannan (LAM) of Mtb which shares some similar properties to LPS, thus BPI-mediated immune response to Mtb can be detectable in TB patients (*41*). BPI has regulatory roles including induction of apoptosis, inhibition of angiogenesis, and lipid metabolism (*42*). These two biomarkers indicate increased inflammation in TB patients with DM. However, mechanisms underlying altered immune responses in DM require further investigation.

There were some limitations. The melioidosis cohort was recruited from a single centre study which may not be representative of all melioidosis patients worldwide. The study site is a tertiary hospital, where melioidosis patients were transferred from adjacent provinces resulting in delayed enrolment, diagnosis, and treatment. Due to a lack of information on interventions or treatments received prior to enrolment into the studies, the impact of drugs on the observed transcriptomic profiles is unknown. Participants were enrolled by the time of established disease with a median of 5 days after hospital admission due to the use of positive culture of Bp as inclusion criteria. Therefore, early transcriptomic responses adjacent to disease onset were not captured. Most patients with melioidosis had one or more co-morbidities that may be confounding factors to delineate the transcriptomic profile associated with DM. Finally, full characterisation of patients’ DM status including Type 1, Type II or other, the length of the disease, as well as treatment was not available in this study, and these features may influence the outcome of this study.

In summary, both melioidosis and TB patients with DM exhibit increased inflammatory responses compared to those without DM, with key dysfunctions of neutrophil function, clotting cascades and platelet function (Figure 5). The reduced interferon immune response in TB patients with DM may be a potential underlying mechanism of increased susceptibility to Mtb infection. In contrast to TB, melioidosis is typically an acute infection, with neutrophil degranulation, coagulation, and platelet activation pathways, more pronounced in melioidosis compared to TB. The degree to which the increased inflammatory response, in particular by neutrophils, is compensatory as a result of uncontrolled infection, or causal for impaired cellular interferon responses by immunomodulation via inflammatory mediators, compromised function of innate immune cells and dysregulation of the adaptive T cell response requires animal models to dissect. Future use of a diabetic mouse model alongside studying early transcriptomic responses to vaccines in people with and without DM will build on our findings. Our findings, across two diseases and five countries, give a general insight into the impact of diabetes on the human immune response to infection, but studies in further diseases and populations will extend the generalisability of our findings.

**Figure 5.**
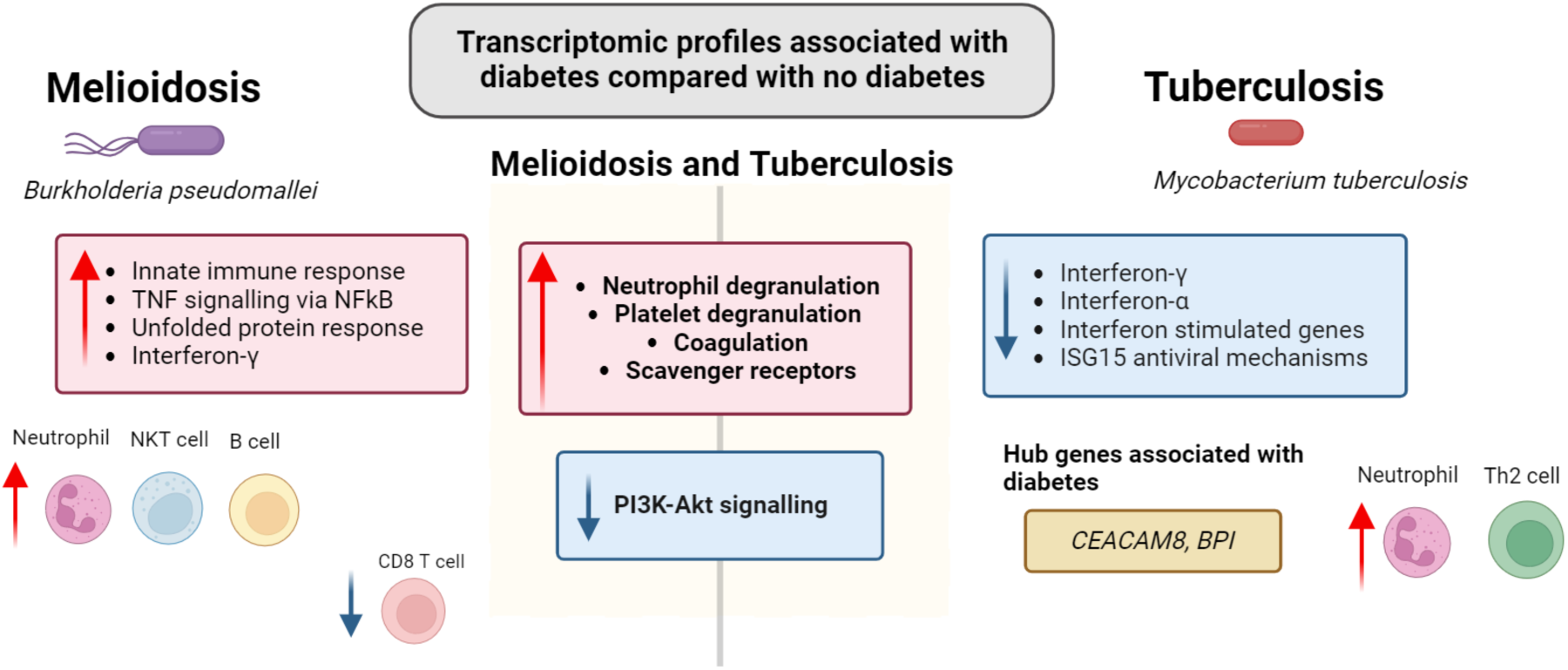
Graphical summary of whole blood transcriptomic profiles associated with increased susceptibility to melioidosis and tuberculosis in people who live with and without diabetes compared with their respective healthy control cohorts. Created with BioRender.com.

## Data Availability

All data produced in the present study are available upon reasonable request to the authors

## Acknowledgements

We thank all the participants and staff at Sunpasitthiprasong Hospital for their laboratory and administrative support. S.J.D. is grateful for the support of a Wellcome Trust Intermediate Clinical Fellowship award ref WT100174/Z/12/Z and is now funded by an NIHR Global Research Professorship ref NIHR300791. We declare no conflict of interest.

## Author contributions

Conceptualization: SJD, JC; Methodology: PR, CE, MA, EM, TEW, SAG; Formal Analysis: PR; Investigation: PR; BK, CE, KJ, SC, PC, MA; Resources: DL, NC, TEW; Writing – Original Draft: PR; Writing – Reviewing & Editing: BK, PC, EM, DL, NC, PK, SJD, TEW, SAG; Supervision: BK, PK, SJD, TEW, SAG; Funding Acquisition: NPJD, DL, SJD

