## Supplementary Figure 1 for "Diabetes mellitus is associated with a shared hyper-inflammatory immune response in melioidosis and tuberculosis patients: an observational case-control study"

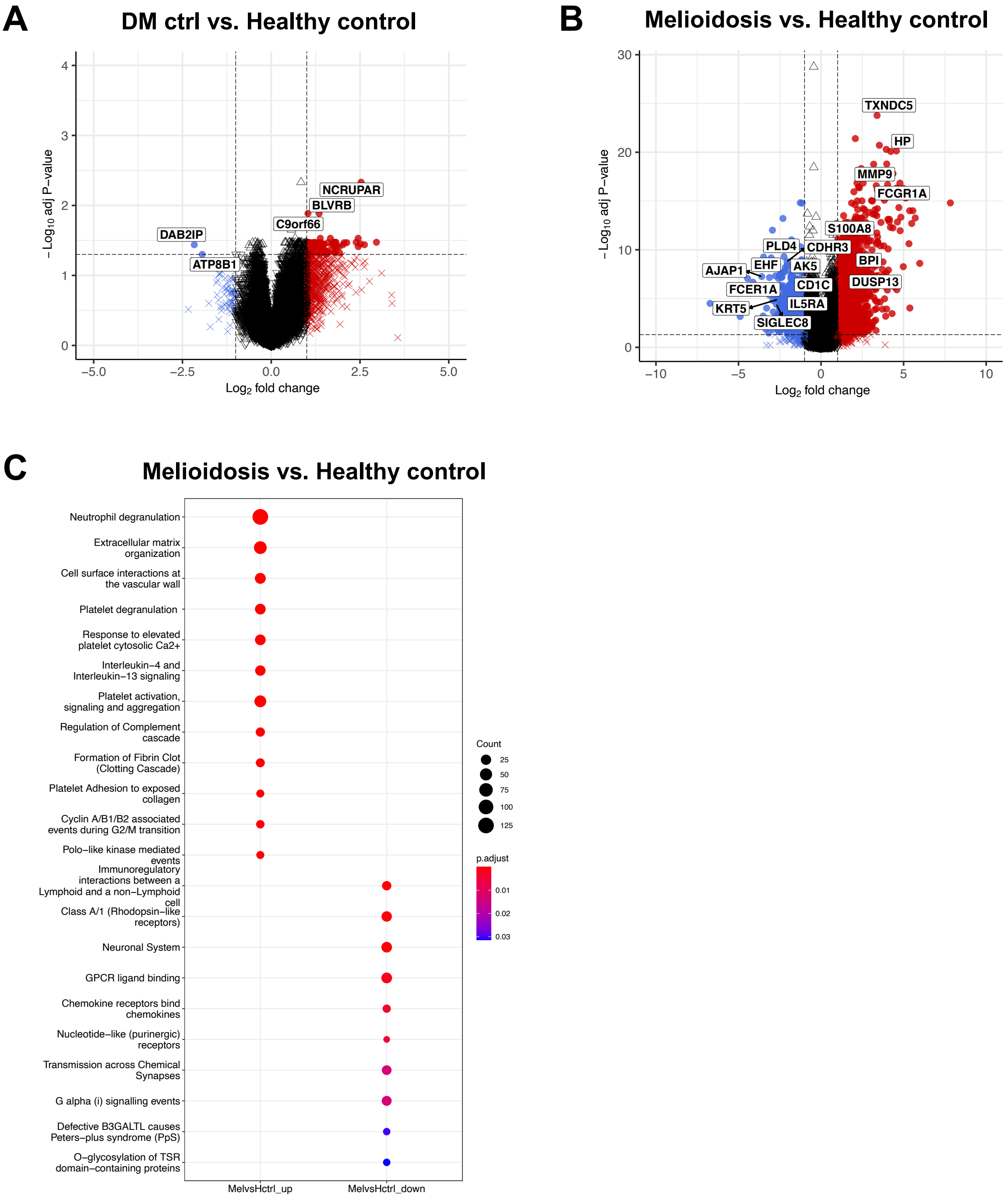


**Supplementary Figure 1. Melioidosis drives widespread immune activation.** (A) Volcano plot of differentially expressed genes between uninfected diabetes (DM) outpatients and healthy donors. (B) Volcano plot of differentially expressed genes between melioidosis patients and uninfected control cohort. Differentially expressed genes were based on absolute (Log$_2$

fold-change)≥ 1 (x-axis) and adjusted P-value < 0.05 (y-axis) (dotted lines). (C) Functional pathway analysis based on Reactome gene sets following differential gene expression (DGE) analysis between melioidosis patients and uninfected healthy donors. The gradient colour bar corresponds to the adjusted P-value. The size of each term is indicated by representative counts (number of DEGs). Differentially expressed genes were pre-filtered based on a cut-off of absolute[Log2 fold-change] ≥1 and adjusted P-value <0.05. MelvsHctrl_up and MelvsHctrl_down = up and downregulated pathways derived from DGE analysis between melioidosis patients compared to uninfected healthy donors respectively.
