## Supplementary Figure 2 for "Diabetes mellitus is associated with a shared hyper-inflammatory immune response in melioidosis and tuberculosis patients: an observational case-control study"

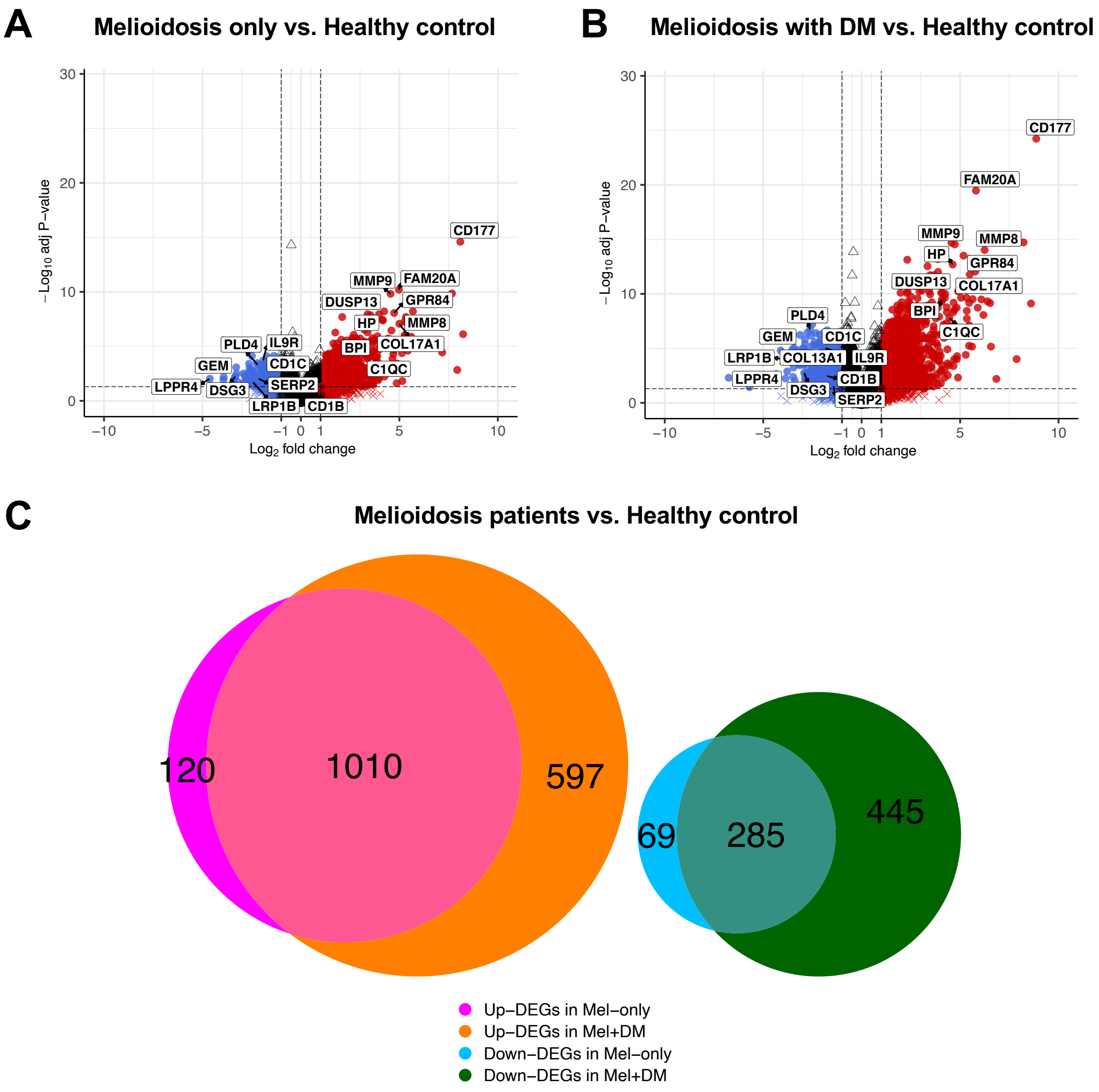


**Supplementary Figure 2. Higher magnitude of transcriptomic response in melioidosis patients with DM.** (A) Volcano plot of differentially expressed genes between melioidosis patients without diabetes and healthy donors. (B) Volcano plot of differentially expressed genes between melioidosis patients with and healthy donors. Differentially expressed genes (DEGs) were based on absolute ($Log_2$ fold-change)≥ 1 (x-axis) and adjusted P-value < 0.05 (y-axis) (dotted lines). (C) Euler diagram shows overlapped and distinct DEGs derived from DGE analyses among melioidosis patients and healthy donors (A, B). Up-DEGs in Mel-only (magenta) and Down-DEGs in Mel-only (blue) = up and downregulated genes derived from DGE analysis between melioidosis patients without diabetes and healthy donors. Up-DEGs in Mel-only (orange) and Down-DEGs in Mel-only (green) = up and up and downregulated genes derived from DGE analysis between melioidosis patients with diabetes and healthy donors.
