## Supplementary Figure 3 for "Diabetes mellitus is associated with a shared hyper-inflammatory immune response in melioidosis and tuberculosis patients: an observational case-control study"

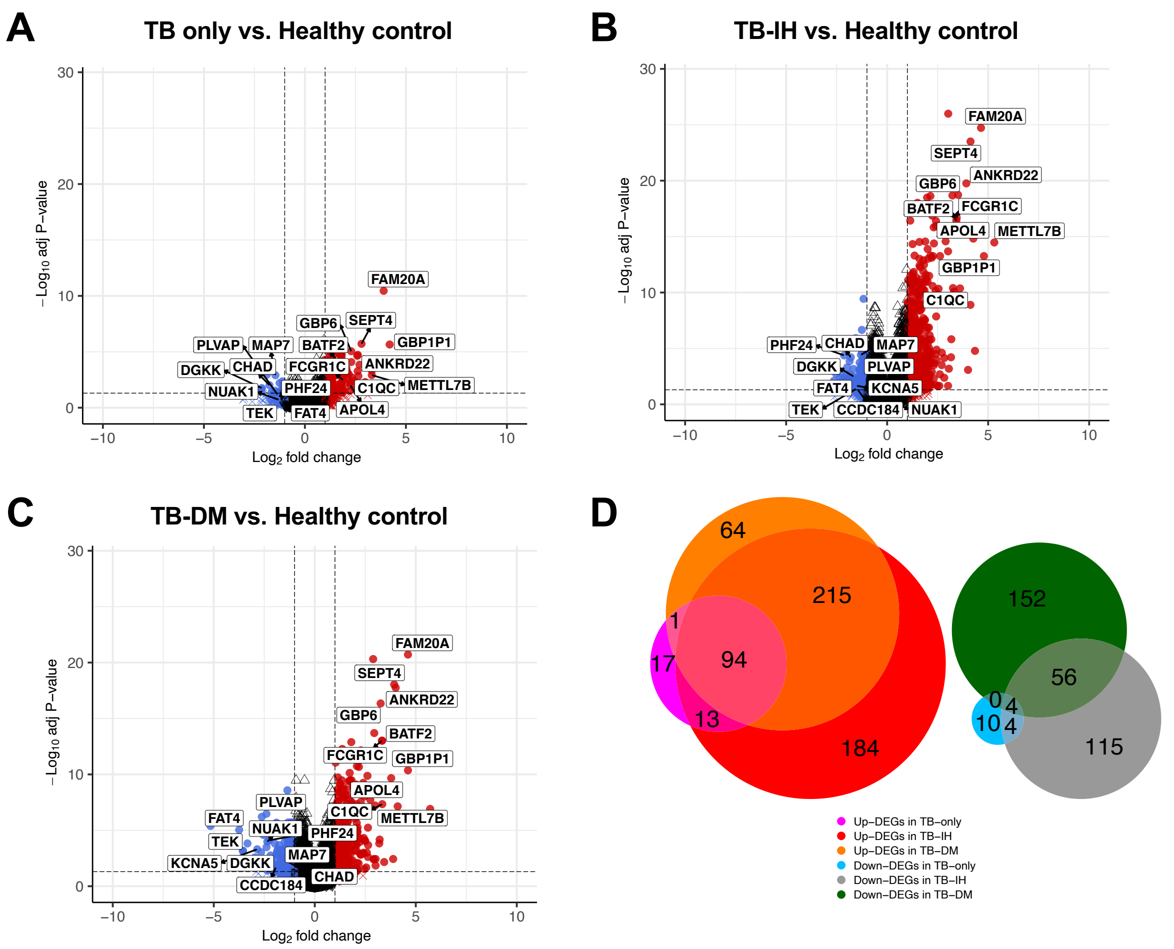


**Supplementary Figure 3. Higher magnitude of transcriptomic responses in tuberculosis patients with intermediate hyperglycaemia and diabetes.** (A) Volcano plot of differentially expressed genes between tuberculosis (TB) patients without diabetes (TB-only) and healthy donors. (B) Volcano plot of differentially expressed genes between TB patients with intermediate hyperglycaemia (TB-IH) and healthy donors. (C) Volcano plot of differentially expressed genes between TB patients with diabetes (TB-DM) and healthy donors. Differentially expressed genes (DEGs) were based on absolute ($Log_2$ fold-change)≥ 1 (x-axis) and adjusted P-value < 0.05 (y-axis) (dotted lines). (D) Euler diagram shows overlapped and distinct DEGs derived from DGE analyses among TB patients and healthy donors (A-C). Up-DEGs in TB-only (magenta) and Down-DEGs in TB-only (blue) = up and downregulated genes derived from DGE analysis between TB patients without intermediate hyperglycaemia and healthy donors. Up-DEGs in Mel-IH (red) and Down-DEGs in Mel-IH (grey) = up and downregulated genes derived from DGE analysis between TB patients with diabetes and healthy donors. Up-DEGs in Mel-DM (orange) and Down-DEGs in Mel-DM (green) = up and downregulated genes derived from DGE analysis between TB patients with diabetes and healthy donors.
