## Supplementary Figure 4 for "Diabetes mellitus is associated with a shared hyper-inflammatory immune response in melioidosis and tuberculosis patients: an observational case-control study"

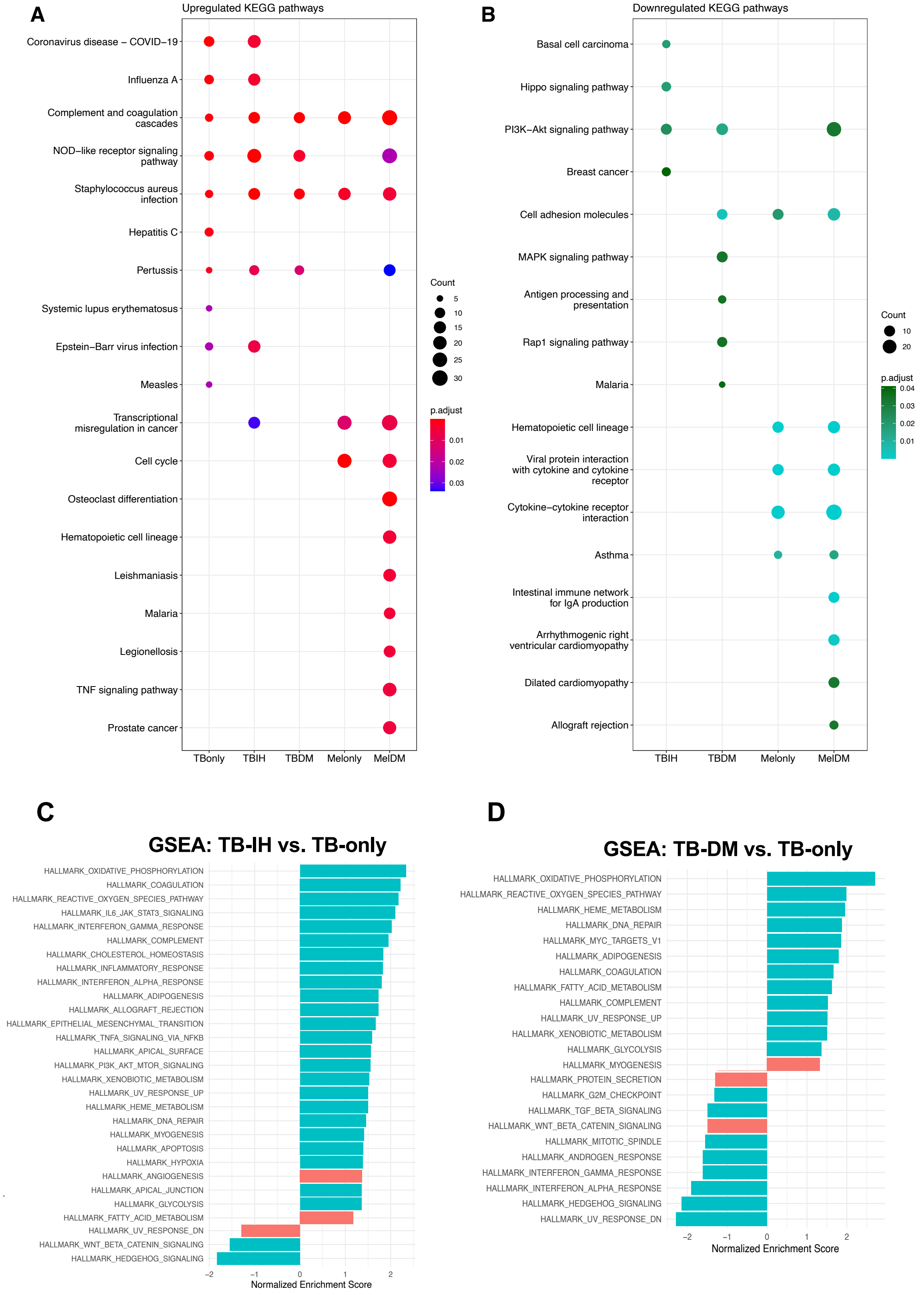
A
Upregulated KEGG pathways
Coronavirus disease − COVID−19
Influenza A
Complement and coagulation cascades
NOD−like receptor signaling pathway
Staphylococcus aureus infection
Hepatitis C
Pertussis
Systemic lupus erythematosus
Epstein−Barr virus infection
Measles
Transcriptional misregulation in cancer
Cell cycle
Osteoclast differentiation
Hematopoietic cell lineage
Leishmaniasis
Malaria
Legionellosis
TNF signaling pathway
Prostate cancer
TBonly
TBIH
TBDM
Melonly
MelDM
Count
5
10
15
20
25
30
p.adjust
0.01
0.02
0.03
B
Downregulated KEGG pathways
Basal cell carcinoma
Hippo signaling pathway
PI3K−Akt signaling pathway
Breast cancer
Cell adhesion molecules
MAPK signaling pathway
Antigen processing and presentation
Rap1 signaling pathway
Malaria
Hematopoietic cell lineage
Viral protein interaction with cytokine and cytokine receptor
Cytokine−cytokine receptor interaction
Asthma
Intestinal immune network for IgA production
Arrhythmogenic right ventricular cardiomyopathy
Dilated cardiomyopathy
Allograft rejection
TBIH
TBDM
Melonly
MelDM
Count
10
20
p.adjust
0.04
0.03
0.02
0.01
C
GSEA: TB-IH vs. TB-only
HALLMARK_OXIDATIVE_PHOSPHORYLATION
HALLMARK_COAGULATION
HALLMARK_REACTIVE_OXYGEN_SPECIES_PATHWAY
HALLMARK_IL6_JAK_STAT3_SIGNALING
HALLMARK_INTERFERON_GAMMA_RESPONSE
HALLMARK_COMPLEMENT
HALLMARK_CHOLESTEROL_HOMEOSTASIS
HALLMARK_INFLAMMATORY_RESPONSE
HALLMARK_INTERFERON_ALPHA_RESPONSE
HALLMARK_ADIPOGENESIS
HALLMARK_ALLOGRAFT_REJECTION
HALLMARK_EPITHELIAL_MESENCHYMAL_TRANSITION
HALLMARK_TNFA_SIGNALING_VIA_NFKB
HALLMARK_APICAL_SURFACE
HALLMARK_PI3K_AKT_MTOR_SIGNALING
HALLMARK_XENOBIOTIC_METABOLISM
HALLMARK_UV_RESPONSE_UP
HALLMARK_HEME_METABOLISM
HALLMARK_DNA_REPAIR
HALLMARK_MYOGENESIS
HALLMARK_APOPTOSIS
HALLMARK_HYPOXIA
HALLMARK_ANGIOGENESIS
HALLMARK_APICAL_JUNCTION
HALLMARK_GLYCOLYSIS
HALLMARK_FATTY_ACID_METABOLISM
HALLMARK_UV_RESPONSE_DN
HALLMARK_WNT_BETA_CATENIN_SIGNALING
HALLMARK_HEDGEHOG_SIGNALING
−2
−1
0
1
2
Normalized Enrichment Score
D
GSEA: TB-DM vs. TB-only
HALLMARK_OXIDATIVE_PHOSPHORYLATION
HALLMARK_REACTIVE_OXYGEN_SPECIES_PATHWAY
HALLMARK_HEME_METABOLISM
HALLMARK_DNA_REPAIR
HALLMARK_MYC_TARGETS_V1
HALLMARK_ADIPOGENESIS
HALLMARK_COAGULATION
HALLMARK_FATTY_ACID_METABOLISM
HALLMARK_COMPLEMENT
HALLMARK_UV_RESPONSE_UP
HALLMARK_XENOBIOTIC_METABOLISM
HALLMARK_GLYCOLYSIS
HALLMARK_MYOGENESIS
HALLMARK_PROTEIN_SECRETION
HALLMARK_G2M_CHECKPOINT
HALLMARK_TGF_BETA_SIGNALING
HALLMARK_WNT_BETA_CATENIN_SIGNALING
HALLMARK_MITOTIC_SPINDLE
HALLMARK_ANDROGEN_RESPONSE
HALLMARK_INTERFERON_GAMMA_RESPONSE
HALLMARK_INTERFERON_ALPHA_RESPONSE
HALLMARK_HEDGEHOG_SIGNALING
HALLMARK_UV_RESPONSE_DN
−2
−1
0
1
2
Normalized Enrichment Score

**Supplementary Figure 4. Increased inflammatory immune responses in melioidosis and tuberculosis patients with diabetes.** Functional pathway analysis based on KEGG gene sets following differential gene expression (DGE) analysis between melioidosis patients compared to uninfected healthy donors and tuberculosis (TB) patients compared to uninfected healthy donors. (A) Up-regulated KEGG pathways derived from the DGE analyses among TB and melioidosis patients compared to their respective uninfected healthy control cohorts. (B) Down-regulated KEGG pathways derived from the DGE analyses among TB and melioidosis patients compared to their respective uninfected healthy control cohorts. The gradient colour bar corresponds to the adjusted P-value. The size of each term is indicated by representative counts (number of DEGs). Differentially expressed genes were pre-filtered based on a cut-off of absolute[Log2 fold-change] ≥1 and adjusted P-value <0.05. Gene set enrichment analysis based on Hallmark gene sets following differential gene expression (DGE) analysis among tuberculosis patients across four study sites. (C) TB patients with intermediate hyperglycaemia (TB-IH, n=44) compared to TB patients without DM (TB-only, n=46). (D) TB patients with diabetes (TB-DM, n=61) compared to TB patients without DM (TB-only, n=46). Normalised enrichment scores are displayed, in which pathways were deemed significant when adjusted P-value <0.05.
