## Supplementary Figure 5 for "Diabetes mellitus is associated with a shared hyper-inflammatory immune response in melioidosis and tuberculosis patients: an observational case-control study"

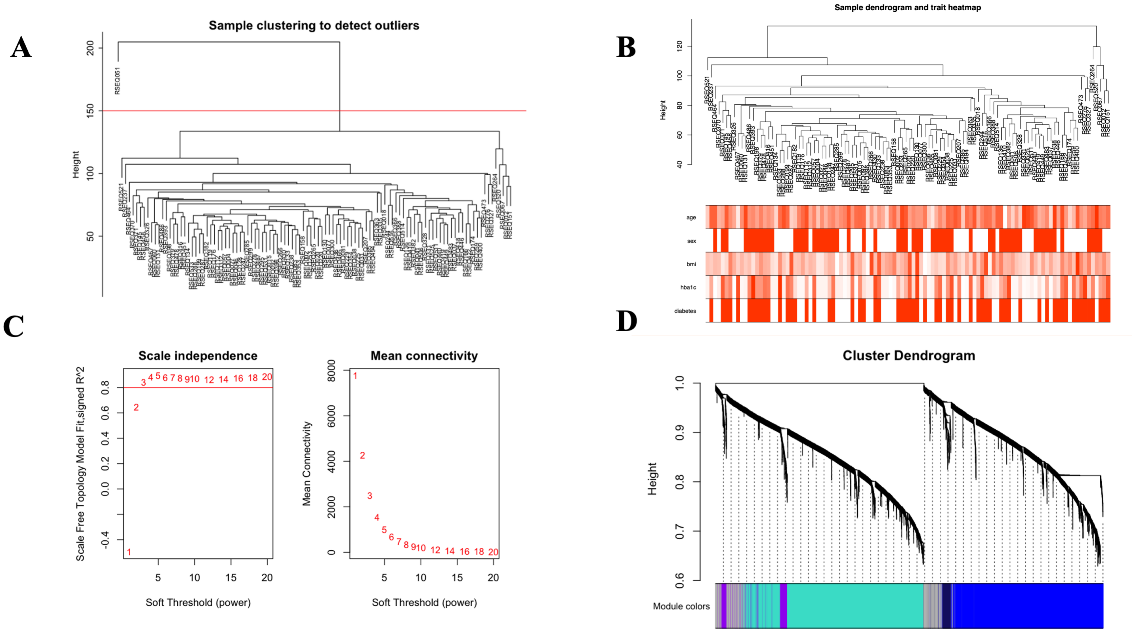


**Supplementary Figure 5. Increased inflammatory immune responses in tuberculosis patients with diabetes.**

Weighted gene co-expression network analysis (WGCNA) was performed in 107 tuberculosis (TB) patients including 61 TB patients with diabetes (DM) and 46 TB patients without diabetes. (A) Sample dendrogram identified and removed one outlier. (B) Sample dendrogram with corresponding clinical data after removing the outlier. (C) Scale-free topological analysis, with scale independence plot indicates scale free topology model fit ($R^2$) and mean connectivity plot with soft threshold on x-axis. (D) Cluster dendrogram of co-expressed module (module eigengene, ME).
