## Supplementary Figure 6 for "Diabetes mellitus is associated with a shared hyper-inflammatory immune response in melioidosis and tuberculosis patients: an observational case-control study"

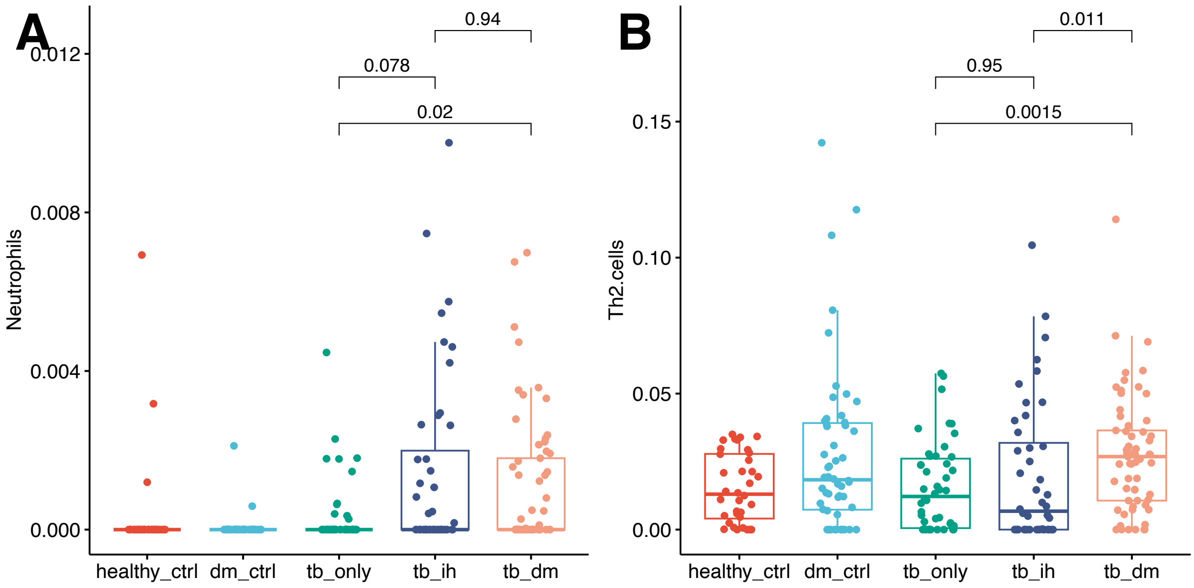


**Supplementary Figure 6**. **Enriched neutrophils and T helper 2 cells were associated with diabetes during tuberculosis.** (A, B) Enrichment of neutrophils and T helper 2 (Th2) cells between tuberculosis patients with diabetes and without diabetes respectively. Cell enrichment was performed using xCell deconvolution method. The statistical analysis was performed using Mann-Whitney test, and the corresponding P-value was displayed on each plot along with median and inter-quartile range boxes.
